# Evaluating generative artificial intelligence’s limitations in health policy identification and interpretation

**DOI:** 10.1101/2024.10.02.24314805

**Authors:** Rory Wilson, Ciara M. Weets, Amanda Rosner, Rebecca Katz

## Abstract

Policy epidemiology utilizes human subject-matter experts (SMEs) to systematically surface, analyze, and categorize legally-enforceable policies. The Analysis and Mapping of Policies for Emerging Infectious Diseases project systematically collects and assesses health-related policies from all United Nations Member States. The recent proliferation of generative artificial intelligence (GAI) tools powered by large language models have led to suggestions that such technologies be incorporated into our project and similar research efforts to decrease the human resources required. To test the accuracy and precision of GAI in identifying and interpreting health policies, we designed a study to systematically assess the responses produced by a GAI tool versus those produced by a SME.

We used two validated policy datasets, on emergency and childhood vaccination policy and quarantine and isolation policy in each United Nations Member State. We found that the SME and GAI tool were concordant 78.09% and 67.01% of the time respectively. It also significantly hastened the data collection processes.

However, our analysis of non-concordant results revealed systematic inaccuracies and imprecision across different World Health Organization regions. Regarding vaccination, over 50% of countries in the African, Southeast Asian, and Eastern Mediterranean regions were inaccurately represented in GAI responses. This trend was similar for quarantine and isolation, with the African and Eastern Mediterranean regions least concordant. Furthermore, GAI responses only provided laws or information missed by the SME 2.14% and 2.48% of the time for the vaccination dataset and for the quarantine and isolation dataset, respectively. Notably, the GAI was least concordant with the SME when tasked with policy interpretation.

These results suggest that GAI tools require further development to accurately identify policies across diverse global regions and interpret context-specific information. However, we found that GAI is a useful tool for quality assurance and quality control processes in health policy identification.

## Introduction

The Analysis and Mapping of Policies for Emerging Infectious Diseases (AMP EID) project employs a standardized protocol to systematically surface, analyze, and categorize health-related, legally-enforceable policies from all United Nations (UN) Member States.^1,2,3,4^ The advent and proliferation of generative artificial intelligence (GAI) technology has created tools that rapidly sift through the wealth of digitized knowledge. This has led to suggestions from affiliates that automating the AMP EID protocol could exponentially decrease the human-hours required to complete the work. We previously explored using GAI tools for this work, but we found that they lacked precision and accuracy when answering questions on nascent research areas. We were also concerned about their relative accuracy in global south countries.

However, GAI tools are becoming increasingly more accurate.^5^ Therefore, we designed a study to understand the extent to which a popular research GAI tool could appropriately identify and interpret relevant policies. We achieved this by systematically assessing the results and sources returned by the system against two validated policy datasets produced by the AMP EID research team.

### Methodology

We used the Default model of Perplexity Pro, produced by Perplexity AI. This answer engine combines traditional search pipelines with large language models (LLMs) produced by integration with Azure OpenAI Service to construct conversational responses to queries.^6^ We chose Perplexity Pro as the GAI tool for this project because it included citations in produced responses. Citations enhanced transparency and facilitated assessments of the sources used to construct responses.

We used two datasets of policies collected by our research team of subject-matter experts (SMEs) utilizing our standardized AMP EID data collection protocol (Supplementary Material) as the standard for tool assessment. Airtable, a cloud based relational database, was used for data collection, while R was used to perform analysis of results.

In order to determine the optimal usage of the GAI tool for our purposes, we created two different query approaches for answering relevant questions for our datasets (See Table 1). However, across both trials, we used an identical protocol for identifying and coding results. We used ‘law’ instead of policy in our queries as the GAI tool was significantly less accurate when the term legally-enforceable policy was used. Terms were entered verbatim into the query line of the GAI tool in numerical order. Searches were performed until a specific policy was identified by the GAI tool or until search terms were exhausted.

**Table 1.** Query terms for identifying relevant vaccination policies as entered into the GAI tool.

| Topic | Order | Query |
| --- | --- | --- |
| Routine Childhood Vaccination | 1 | Is there a law that allows the government of [Country] to mandate that a child receives a routine vaccination? |
|  | 2 | What law allows the government of [Country] to require routine immunizations? |
|  | 3 | Is there a legally-enforceable mandate in [Country] for children to receive routine vaccinations? |
| Emergency Vaccination | 1 | In the case of a public health emergency, can the government of [Country] mandate that citizens receive a compulsory vaccination? |
|  | 2 | What law allows the government of [Country] to require vaccinations for citizens during an emergency? |
|  | 3 | What law gives the government of [Country] emergency powers? |

We first used our vaccination dataset to determine the ability of the GAI tool to accurately identify the most relevant laws for routine childhood and emergency vaccination of the population. We then used our quarantine and isolation dataset to understand if and how the GAI tool could identify and interpret relevant laws when given specific parameters. The protocols for each arm of the project are detailed below. Democratic People’s Republic of Korea was excluded from the study, as there was not enough publicly available information to verify the GAI result.

For all searches, results and citations were reviewed to ensure that the tool was using information from reputable sources and was not citing work previously published as part of the AMP EID project. If the tool was sourcing from unreliable materials, the query was rerun with the addition of the sentence, “Use only peer-reviewed sources when producing a response.” In the case that work related to AMP EID was cited, the query was rerun with a request to exclude information specifically from the AMP EID-related resource. In the quarantine and isolation dataset, whether or not this new answer was significantly different from the answer including AMP EID was noted.

Concordance was calculated as the number of times GAI organically produced the same answer to the queries as the research team of subject-matter experts (SMEs). This was calculated as:

Concordance rate = ((# entries coded as Condordance = “Yes”)/(Total # entries)) *100

#### Language Analysis

In order to identify biases of the GAI tool in surfacing and interpreting policies written in languages other than English, we began by identifying countries that utilize any of the 6 official UN languages as an “official language”.⁷ We then used our master repository of policies included in the AMP EID database (2,905 policy documents) to identify which languages each country uses to publish policies. For nations that have multiple official languages, yet in practice use only one language to write policies, we filtered out those official languages that are not used empirically. We then utilized the concordance rate calculation to determine the fidelity of the GAI tool query responses to those of the SME research team in each language.

#### Vaccination Dataset

For each UN Member State, a series of questions were systematically entered into the query line for routine childhood vaccinations and emergency vaccination (See Table 1; Figure 1). All queries were entered only in English. If the policy included in the verified database was not surfaced through queries, we asked for the policy by name in the original language, using the convention, “Answer the query by searching for [name of policy *in original language*].” After exhausting the query protocol, findings were coded according to surfaced results (Figure 1).

**Figure 1.**
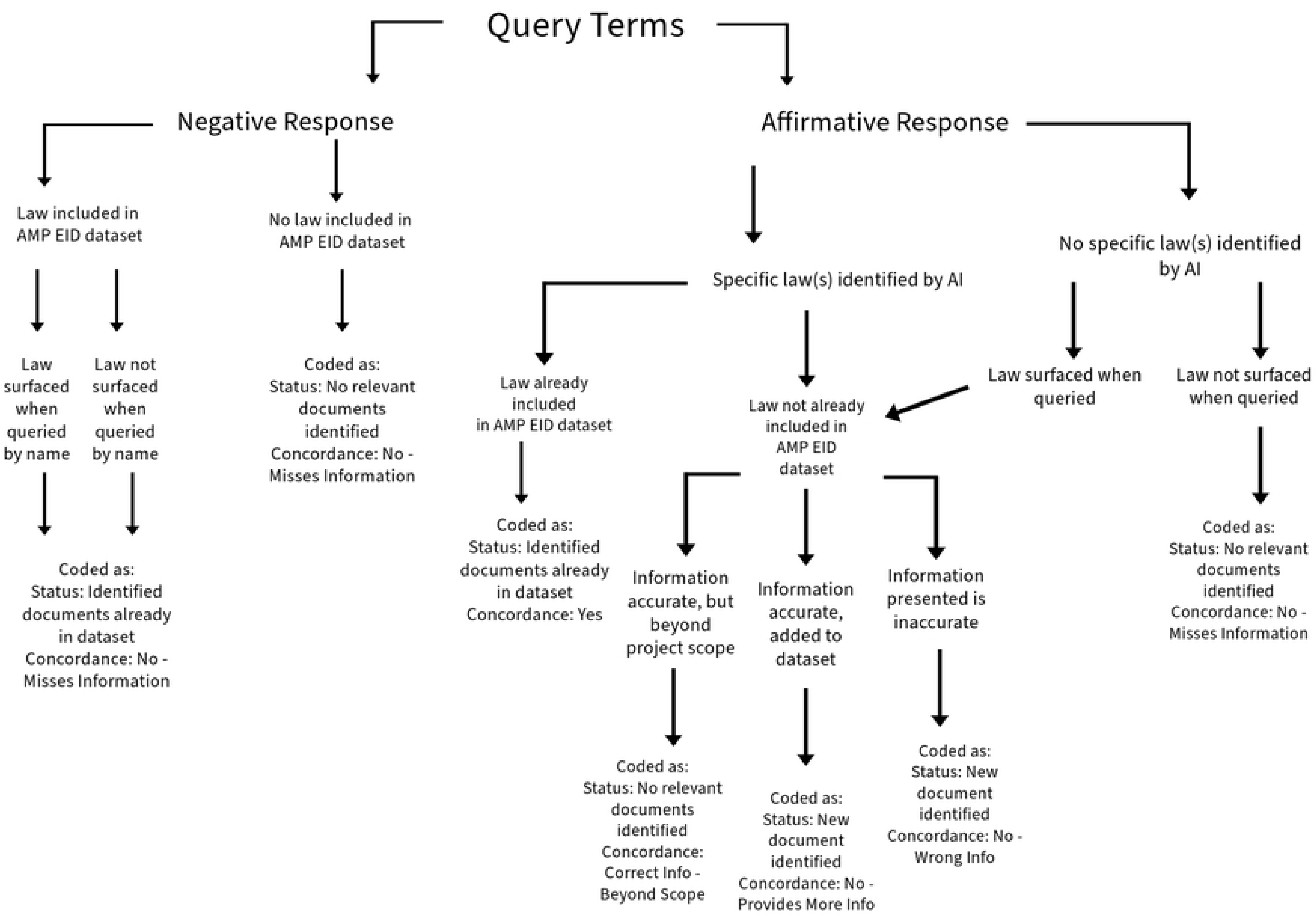
Decision and coding tree for Vaccination Methodology. This decision tree, read top to bottom, was used across all UN Member States. For each country, query terms were used, and, after exhausting all query terms, the aggregate responses were used to make decisions according to this standardized tree. All possible responses result in a coding directive, which are color coded at the base of the tree.

#### Quarantine and Isolation Dataset

For each UN Member State, a series of questions were systematically entered as one search thread into the query line for quarantine and isolation (See Table 2; Figure 2). Policies pertaining to borders and international travelers were specifically excluded. If these were surfaced, the query was rerun with modifications to exclude them. Furthermore, the term ‘isolation of contacts’ was used as a proxy for quarantine in question 5 to help filter out quarantine policies pertaining to international borders and any maritime laws. Once the correct policies were identified, the term “quarantine” is used from question 6 onwards. Specific COVID-19 policies were also excluded unless the country had no other non COVID-19 policies previously identified by the SME. After exhausting the query protocol, findings were coded according to surfaced results (Figure 3).

**Table 2.** Query terms for identifying relevant quarantine and isolation policies as entered into the GAI tool, including question modifications to be entered if the GAI response meets the conditions included in A or B.

| Topic | Order | Query and Modifications |
| --- | --- | --- |
| Isolation | 1 | What law allows the government to isolate sick people in [Country]?<br>A. Modified query if the response is about border:<br>(i) Excluding laws about borders and travelers, what law allows the government to isolate sick people in [Country]?<br>B. Modified query if the response is about COVID-19:<br>(i) Excluding legal responses to COVID-19, what law allows the government to isolate sick people in [Country]? |
|  | 2 | In this law (or these laws), what level of government has the authority to isolate sick people? |
|  | 3 | Does the law have any enforcement mechanisms or penalties if someone violates isolation? |
|  | 4 | Does the law limit isolation to a list of diseases?<br>A. Subsequent query if isolation is limited to a list of diseases, but diseases are not mentioned in response:<br>i. What are these diseases? |
| Quarantine | 5 | Does this law allow the government to isolate contacts of infectious disease? |
|  | 6 | In this law, what level of government has the authority to quarantine contacts of infectious disease? |
|  | 7 | Does the law have any enforcement mechanisms or penalties if someone violates quarantine? [if a contact violates isolation] |
|  | 8 | Does the law limit quarantine [the isolation of contacts] to a list of diseases?<br>A. Subsequent query if quarantine is limited to a list of diseases, but diseases are not mentioned in response:<br>i. What are these diseases? |

**Figure 2.**
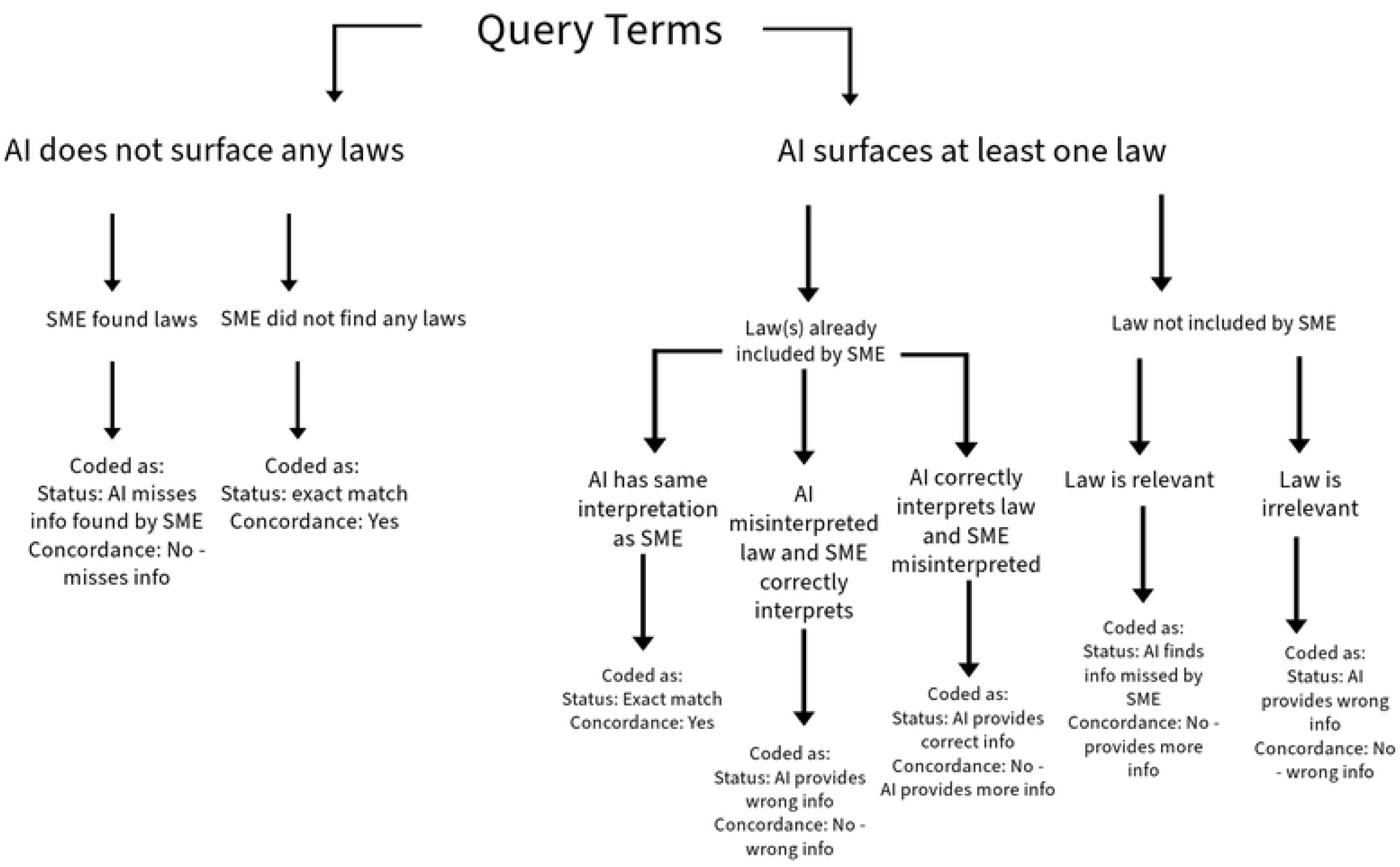
Decision and coding tree for quarantine and isolation law identification and interpretation. This decision tree, read top to bottom, was used across all UN Member States. For each country, query terms were used, and, after exhausting all query terms, the aggregate responses were used to make decisions according to this standardized tree. All possible responses result in a coding directive.

**Figure 3.**
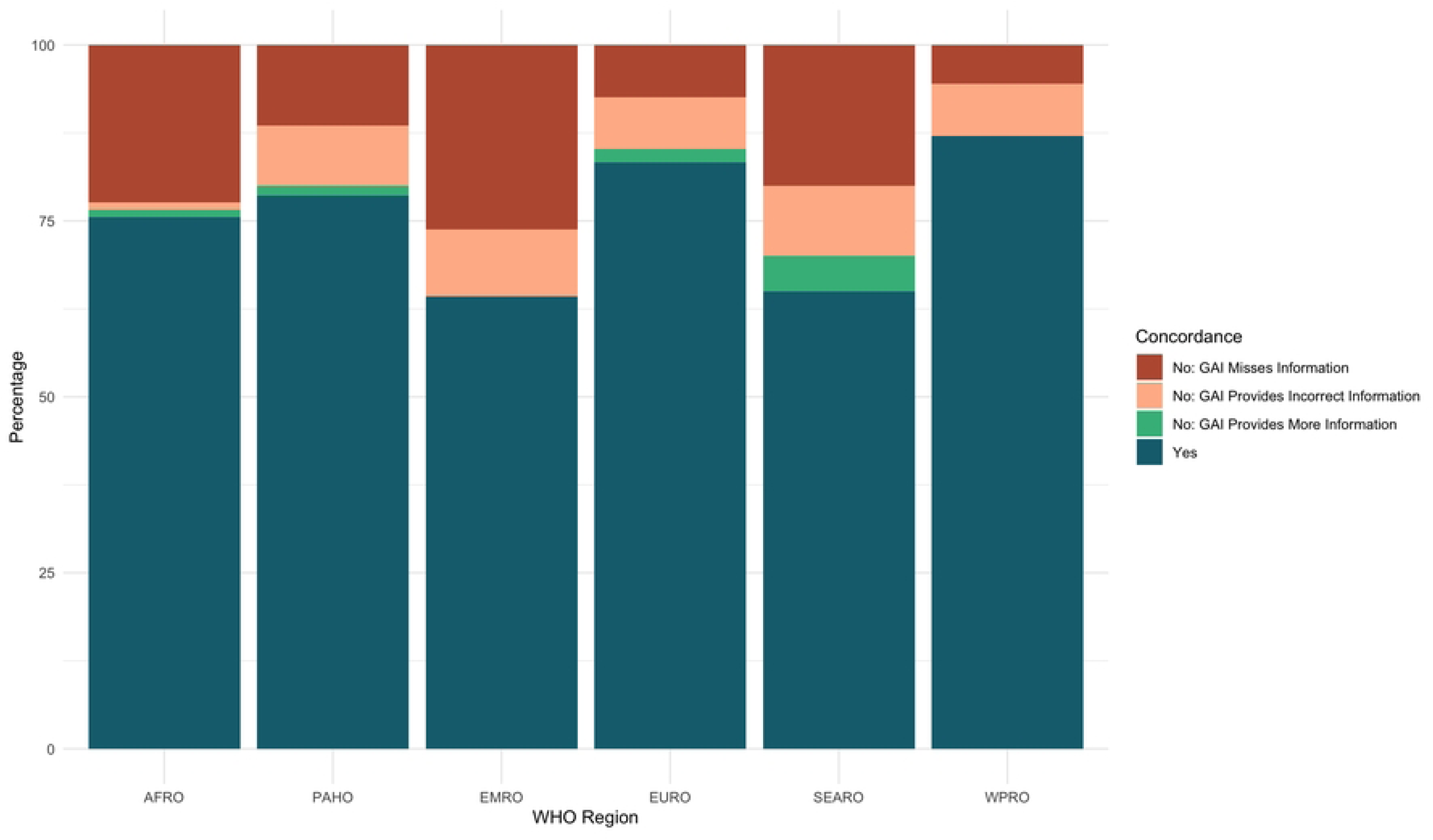
Unfiltered vaccination concordance rates per WHO region.

## Results

### Vaccination

For the vaccination dataset, the methodology asked the GAI tool whether or not there was a legally-enforceable routine childhood vaccination mandate or emergency powers for mandatory vaccination of the domestic population during a crisis. When asked this binary question, the concordance rate between the GAI tool and the human research team was found to be 78.09% (302/388 responses). We filtered out the countries for which the research team and the GAI tool found that there is no universal legal mandate for vaccination, thus isolating the search only to countries for which the research team or the GAI tool had independently found that relevant policies did exist resulting in the concordance rate dropping to 63.20% (146/231 responses).

Concordance was not evenly distributed across World Health Organization (WHO) Regions. When considering the complete, unfiltered dataset, responses on countries within the Western Pacific (WPRO) and European (EURO) regions were the most concordant with 87.04% and 83.33% concordance respectively. Responses from the GAI tool on the presence or absence of vaccination laws in the American (PAHO) and African (AFRO) regions were found to be in agreement with that of the research group for between 78.57% and 75.53% of entries.

Due to the number of states that are documented to lack a legal requirement for routine or emergency vaccination, the inclusion of these countries obfuscates information on the ability of the GAI tool to accurately retrieve policy information across WHO regions. Upon filtering out countries for which there was concordance between the research team and the GAI tool on the lack of legal vaccination requirements, greater diversity in the accuracy of the tool across regions appeared. The filtration process removed 157 responses (40.46%) from the original dataset, leaving 231 responses. While many of the general spatial trends held, the concordance rate fell across regions. WPRO, EURO, and PAHO remained most accurately represented WHO regions with a respective concordance rate of 75.00%, 73.53%, and 71.71%. By contrast, countries in the AFRO, SEARO and EMRO regions were inaccurately represented by the GAI tool over half of the time. Responses from the GAI tool for countries in EMRO region were in concordance with the research team 46.43% of the time, while responses for countries in the AFRO region were in concordance for 45.24% of entries and responses for SEARO countries were in concordance only 41.67% of the time (Figure 4). The significant gap between the concordance rate in the two groups of three countries is stark and notable. Countries in the South-East Asian (SEARO) and Eastern Mediterranean (EMRO) region were the least accurately represented, with concordance rates of 65.00% and 64.29%, respectively (Figure 3).

**Figure 4.**
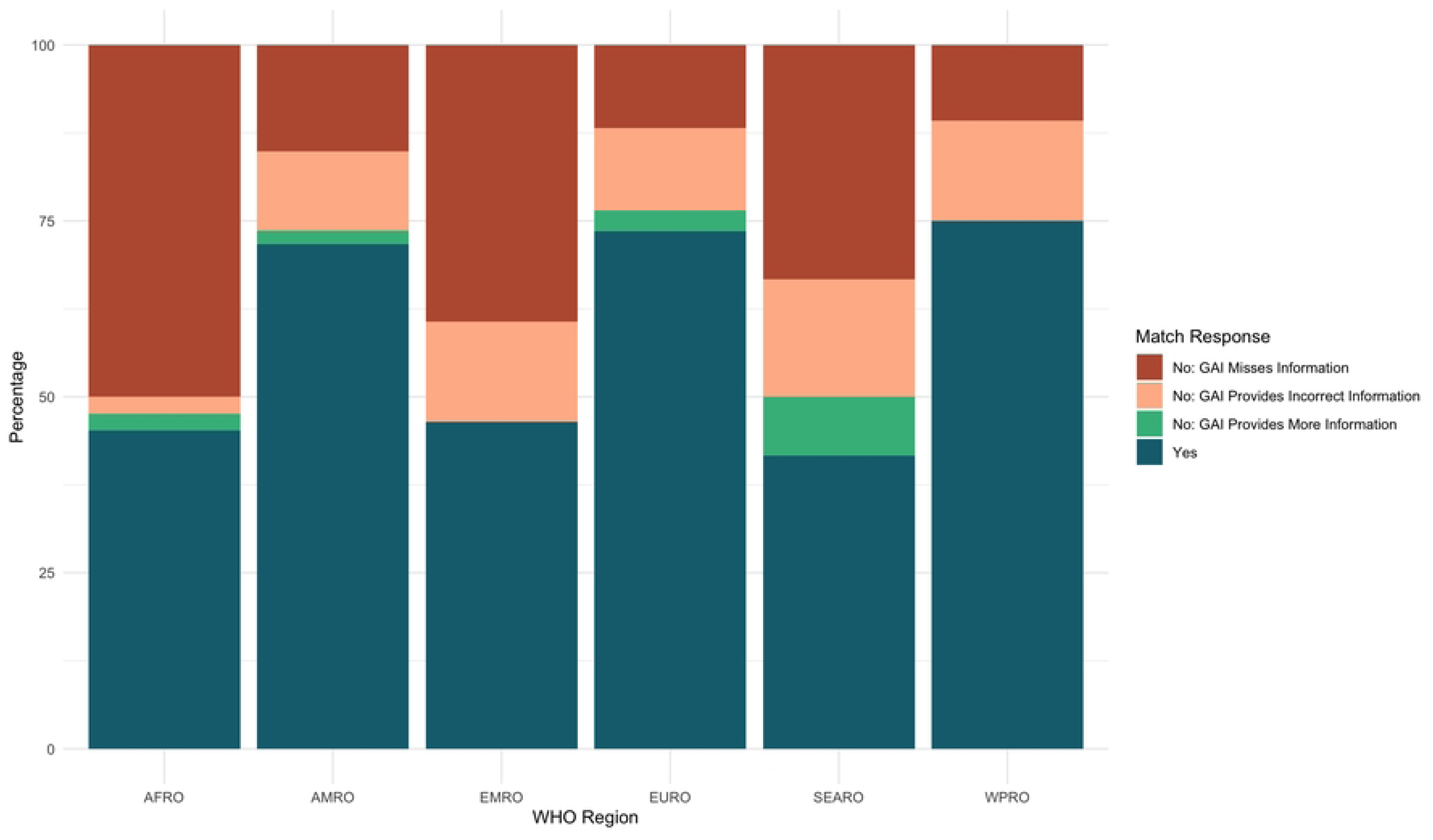
Filtered vaccination concordance rates per WHO region.

For five entries (5/233; 2.14%), the GAI tool identified a policy that had not previously been surfaced by the research team. Of the five instances, one was surfaced through queries about routine childhood vaccinations, while the remaining four were identified through queries pertaining to emergency vaccination.

The concordance for emergency vaccination laws in each UN Member State and concordance for childhood vaccination in each UN Member State is shown in Figure 5.

**Figure 5:**
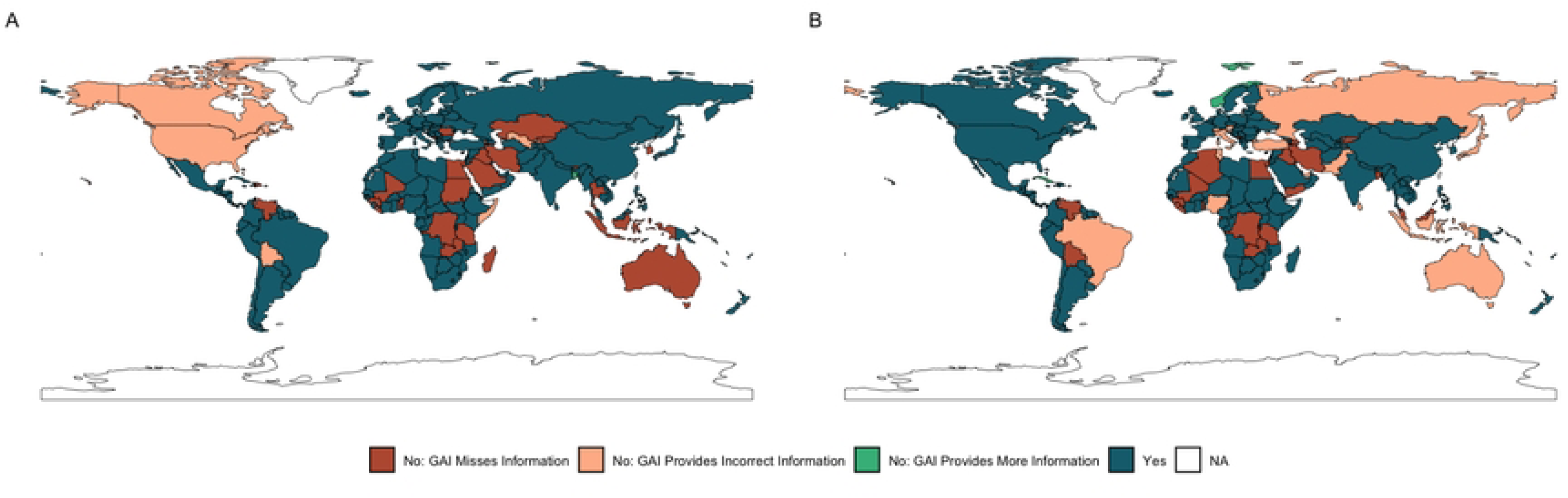
Maps of the concordance between SME research team and GAI tool on routine and emergency vaccination policies in each UN Member State. Panel A includes data on routine childhood vaccination policies, while panel B includes data on emergency powers for vaccination.

### Quarantine and Isolation

For the quarantine and isolation dataset, the methodology asked the GAI tool to surface and interpret any existing policies in the country which allowed for the isolation of sick people and the quarantine of contacts in the domestic population. When asked these successive questions, the concordance rate between the GAI tool and the SME was 67.01% (1040/1552 responses).. For 10 (10/233, 4.29%) countries, temporary COVID-19 policies are used in the absence of standing quarantine and/or isolation authority policies. Their impact on the overall results was statistically insignificant so they were not filtered from our analysis.

Concordance was unevenly distributed across WHO Regions. Quarantine and isolation policies in countries within the WPRO region were the most concordant with 91.67% concordance between the GAI tool and the SME. SEARO, EURO and PAHO regions were moderately concordant with 71.25%, 66.91% and 65.00% concordance respectively. Countries in the EMRO and AFRO regions were the least concordant, with concordance rates of 60.12% and 56.65% respectively (Figure 6).

**Figure 6:**
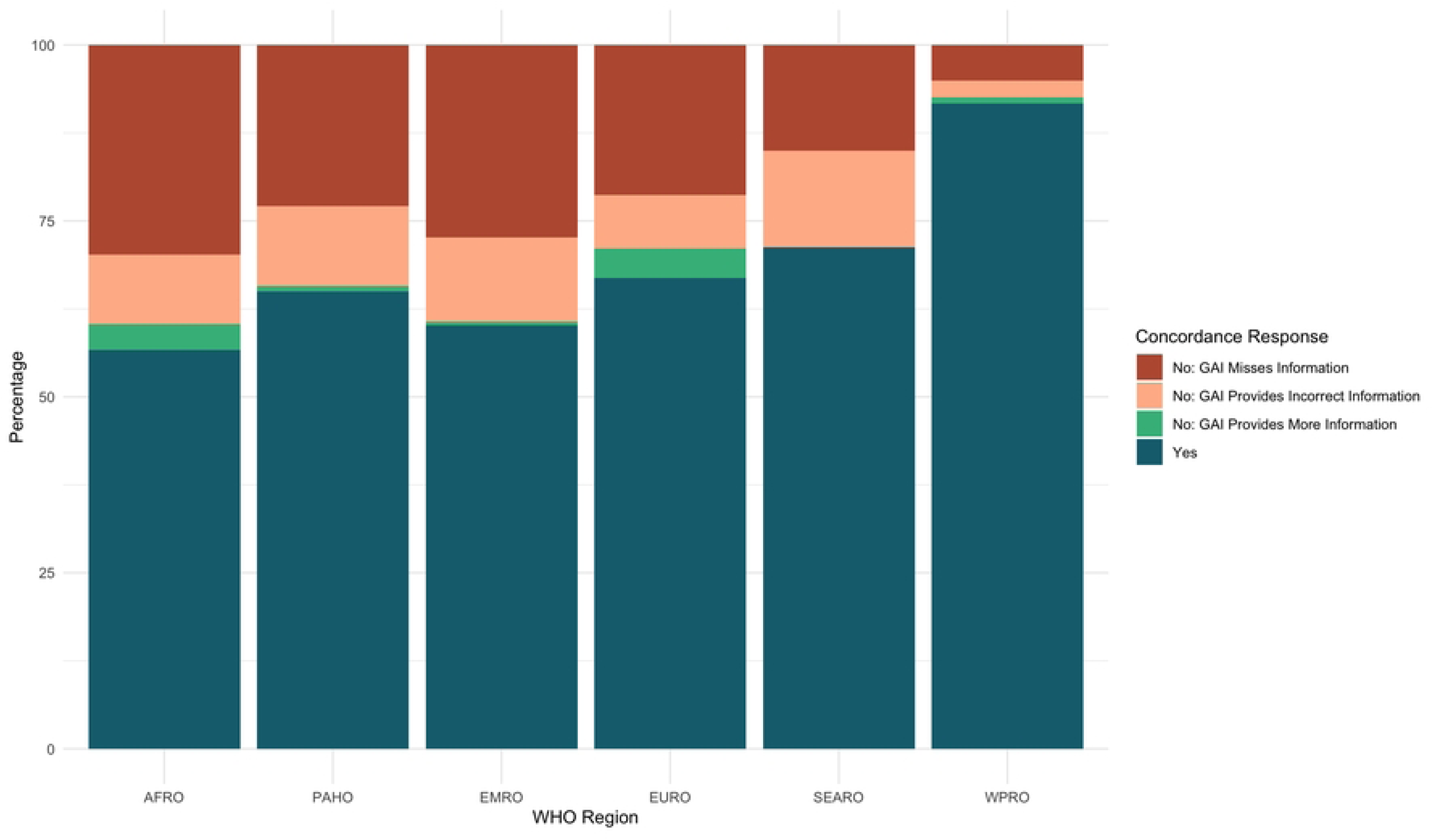
Quarantine and Isolation concordance rates per WHO region.

We suspected that the relatively high rates of concordance in WPRO countries was because a significant number (12/37) use English as an official language, meaning they routinely produce government documents in English. Therefore, we analyzed whether the GAI was better at identifying and interpreting policies in countries with policies written in English than in other languages. We found that the GAI exactly matched the SME results or provided more information 81.56% (398/488 responses) of the time in the 61 countries with policies written in English. In contrast, the GAI only found exact matches to SME or provided more information in 63.86% (697/1064 responses) of the time in the 133 countries which did not have policies written in English.

We then decided to assess the concordance rates for UN Member States which use each of the UN languages as either an official or national language and for which the SME have recorded policies written in these languages. This revealed that countries using Mandarin were the most concordant at 100%, however, this is because only China and Singapore used Mandarin in our dataset. Countries using English were the second most concordant with a rate of 80.12% This was followed by countries using Russian with a concordance of 67.86% and countries using Arabic with a concordance rate of 63.04%. The least concordant countries were countries using Spanish with 57.50% and followed by countries using French with 46.78%.

The overall non-concordance rate between the GAI tool responses and the human research team 32.99% (512/1552 responses), which was broken down into three categories. The GAI missed information found by the SME for 21.71% (337/1552) of total responses, accounting for 65.82% (337/512) of non-concordant responses. The GAI provided wrong information when compared to SME (based on a third reviewer adjudication) for 8.89% (138/1552) of total responses, which accounted for 26.95% (138/512) of the non-concordant responses. Furthermore, the GAI found information which was missed by the SME (based on a third reviewer adjudication) for 2.38% (37/1552) of total responses, accounting for 7.23% (37/512) of non-concordant responses.

Notably, AMP EID was cited as a primary source in 9.13% (139/1552) of the GAI responses. When the search was rerun specifically excluding AMP EID as a source, there was a significant difference in the GAI response 35.25% (49/139) of the time and no significant difference 64.75% (90/139) of the time.

The concordance for the identification of isolation laws (Prompt 1) in each UN Member State is shown in Figure 7 (panel A) whilst the concordance for the identification of quarantine laws (Prompt 6) in each UN Member State is shown in Figure 7 (panel B).

**Figure 7:**
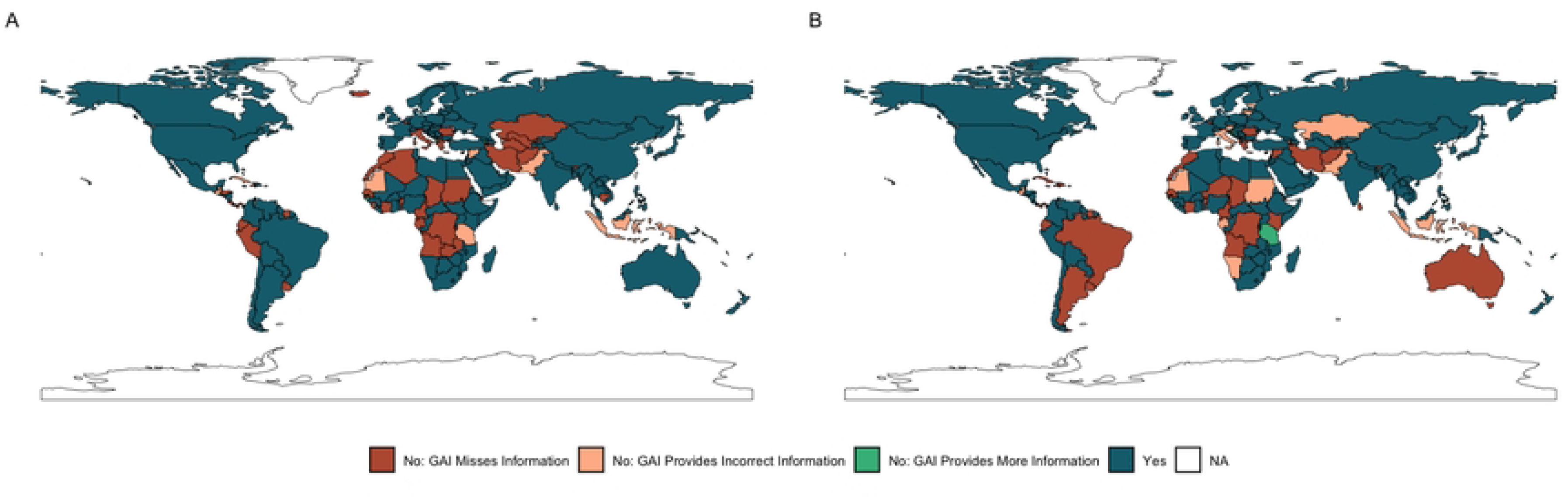
Maps of the concordance between SME research team and GAI tool on quarantine and isolation policies in each UN Member State. Panel A includes data on isolation policies which were surfaced through the first query in the series, while panel B includes data on quarantine policies surfaced by the sixth query of the series.

## Discussion

Despite the GAI tool correctly identifying and interpreting an overall majority of policies in both datasets, it was still significantly non-concordant with the SMEs. Furthermore, the GAI responses only provided laws or information missed by the SME 2.14% and 2.48% of the time for the vaccination dataset and the quarantine and isolation dataset respectively.

Our analysis revealed that the GAI tool was least concordant when identifying and interpreting policies in the AFRO and EMRO WHO regions in both datasets and SEARO for the vaccination dataset. There are likely several reasons for this. There could be linguistic biases causing the lower concordance rates in francophone AFRO countries, as we found French to be the least concordant UN language. Often in these regions, the SME identified policies by searching legal gazettes. The GAI rarely cited legal gazettes which could have contributed to the lack of concordance. Likewise, the GAI was less effective at identifying provisions relevant to health in non-health related policies. For instance, in many EMRO nations, routine childhood vaccination mandates are included in children’s rights and welfare laws, as opposed to being included in public health or infectious disease laws, which is more common in other regions. Thus, the GAI tool’s difficulty with identifying relevant provisions in diverse policies and in languages other than English may account for some of the regional gaps identified in this study. Regardless of the mechanism, the relative inaccuracy of GAI in these critically important regions should offer caution to global health policy researchers on the risk of solely relying on GAI tools.

Within the quarantine and isolation dataset, which assessed the ability of the GAI tool to interpret the contents of policy, as opposed to simply surfacing it, the concordance gap between the GAI tool and SME research team was most notable for queries that required the tool to analyze the policy. This occurred most often when interpreting enforcement mechanisms and disease lists. Importantly, as these prompts required the GAI to conduct a more detailed interpretation of the policies, it raises questions as to the ability of the GAI tool to perform in depth interpretation and policy analysis.

We also encountered issues in generating targeted responses. In quarantine and isolation search threads, the GAI tool exhibited a strong tendency to only refer to COVID-19 policies. This was likely due to the quantity of sources available. However, this required repeated input of exclusion terms by the researcher. For example, nearly all countries required exclusion input for the first quarantine and isolation prompt. The degree of human oversight required to instruct the GAI tool to generate appropriate responses was significant and highlights the risks of inaccuracy when using the GAI tools with limited human oversight.

A common concern when using GAI tools is the risk of hallucination–when AI generates incorrect or misleading results–and we were cognizant of this throughout the study.^8^ We found that within the quarantine and isolation dataset, the GAI tool hallucinated policies that were determined to not exist for Moldova, Italy, and Guatemala. This was based on the fact that there was nothing in the cited supporting evidence referencing these laws nor could the SME find these laws, despite extensive secondary searches.

Despite these inaccuracies and biases, our SMEs ultimately found the GAI tool to be useful for quality assurance and quality control of the identification of vaccination, quarantine, and isolation policies. We believe that the current optimal use for GAI tools in identifying public health policies is as a second reviewer for quality assurance and control of policy identification. However, we did not have confidence in using the tool for interpretation of quarantine and isolation policies. This is in contrast to a previous study comparing GAI and human coders in legal ruling interpretation, which suggested that GAI could be used initially as first reviewer then humans as second reviewer.^7^ The analysis of our GAI tool interpretation responses, lead us to conclude that the current GAI technology is insufficiently developed to reliably interpret these health policies. However, this may change as GAI technology advances over the coming years so we will continuously monitor the evolution of GAI.

There were several limitations in our study. Primarily, we only used one GAI tool. When assessing concordance, we assumed that the SME results are the gold standard. The phrasing of our prompts may have resulted in unintended biases towards inclusion or exclusion of certain laws. Lastly, relying on English language only prompts may have biased the responses against countries which do not have policies written in English.

## Conclusion

We found that GAI is a useful tool to incorporate into quality assurance and quality control for public health policy identification. However, GAI does not yet accurately provide information across diverse global regions and languages, nor does it accurately interpret detailed context-specific information. We suggest that GAI currently should not be relied upon as a primary reviewer in health policy identification or interpretation, but is effective as a second or third reviewer in health policy identification.

## Data Availability

All relevant data is available in a public open access repository. Data can be found on GitHub at https://github.com/cghss/AI_policy_epi

## Acknowledgments

Study was funded using a grant from Rockefeller Foundation. The funder had no part in the conceptualization, design or analysis of the study. The authors would like to acknowledge the assistance of Zahra Izzi in the generation of methodology figures.

## Supporting Information

Analysis and Mapping of Policies for Emerging Infectious Disease (AMP EID) policy identification, preliminary screening, and collection protocol. Information about the AMP EID inclusion criteria development process and data taxonomy are included within this document.

## References

1. Katz R, Graeden E, Kerr J, Eaneff S. Policy Epidemiology: Identifying What Works in Outbreak Preparedness and Response. Health Affairs. 2023 Sep 14. Available from: https://www.healthaffairs.org/content/forefront/policy-epidemiology-identifying-works-outbreak-preparedness-and-response

2. Katz R, Graeden E, Kerr J, Eaneff S. Tracking the flow of policy: Applying a new approach for tracking the flow of health policy. Milbank Q. 2023;101(3):632–652.

3. Weets CM, Katz R. Global approaches to tackling antimicrobial resistance: a comprehensive analysis of water, sanitation and hygiene policies. BMJ Glob Health. 2024;9(2):e013855.

4. Ljungqvist GV, Weets CM, Stevens T, Robertson H, Zimmerman R, Graeden E, et al. Global Patterns in Access and Benefit-Sharing: A Comprehensive Review of National Policies. medRxiv [Preprint]. 2024 Jul 12:2024.07.12.24310347.

5. OpenAI, Achiam J, Adler S, Agarwal S, Ahmad L, Akkaya I, et al. GPT-4 Technical Report. arXiv.. 2023 Mar 15:2303.08774.

6. Perplexity AI. FAQ. Available from: https://www.perplexity.ai/hub/faq?fob=rmseVsxOs82GXAaM

7. United States Central Intelligence Agency. “CIA World Factbook”. 2024. Available from: https://www.cia.gov/the-world-factbook/field/languages/

8. Choi JH. How to use large language models for empirical legal research. J Inst Theor Econ. Minnesota Legal Studies Research Paper No. 23-23. Available from: https://ssrn.com/abstract=4536852

9. Maynez J, Narayan S, Bohnet B, McDonald R. On faithfulness and factuality in abstractive summarization. arXiv:2005.00661. 2020 May 2. Available from: 10.48550/arXiv.2005.00661.

